# Epidemiology and aetiology of moderate to severe diarrhoea in hospitalised HIV-infected patients ≥5 years old in South Africa, 2018-2021: a case-control analysis

**DOI:** 10.1101/2023.02.23.23286353

**Authors:** Siobhan L. Johnstone, Linda Erasmus, Juno Thomas, Michelle J. Groome, Nicolette M. du Plessis, Theunis Avenant, Maryke de Villiers, Nicola A. Page

## Abstract

Diarrhoea is a recognised complication of HIV-infection, yet there are limited local aetiological data in this high-risk group. These data are important for informing public health interventions and updating diagnostic and treatment guidelines. This study aimed to determine the pathogenic causes for diarrhoeal admissions in HIV-infected patients compared to hospital controls between July 2018 and November 2021.

Admitted diarrhoeal cases (n=243) and non-diarrhoeal hospital controls (n=101) ≥5 years of age were enrolled at Kalafong, Mapulaneng and Matikwana hospitals. Stool specimens/rectal swabs were collected and pathogen screening performed on multiple platforms. Differences in pathogen detections between cases and controls, stratified by HIV status, were investigated.

The majority (n=164, 67.5%) of diarrhoeal cases with known HIV status were HIV-infected. Pathogens could be detected in 66.3% (n=228) of specimens, with significantly higher detection in cases compared to controls (72.8% versus 50.5%, *p*<0.001). Amongst HIV-infected participants, prevalence of *Cystoisospora* spp. was significantly higher in cases than controls (17.7% versus 0.0%, *p*=0.028), while *Schistosoma* was detected more often in controls than cases (17.4% versus 2.4%, *p*=0.009). Amongst the HIV-uninfected participants, prevalence of *Shigella* spp., *Salmonella* spp. and *Helicobacter pylori* was significantly higher in cases compared to controls (36.7% versus 12.0%, *p*=0.002; 11.4% versus 0.0%, *p*=0.012; 10.1% versus 0.0%, *p*=0.023).

Diarrhoeal aetiology differed by HIV status, with *Shigella* spp. (36.7%) and *Salmonella* spp. (11.4%) having the highest prevalence amongst HIV-uninfected cases and *Shigella* spp. (18.3%), *Cystoisospora* (17.7%), and *Cryptosporidium* spp. (15.9%) having the highest prevalence amongst HIV-infected cases. These differences should be considered for the development of diagnostic and treatment guidelines.

## Introduction

Diarrhoeal diseases pose a significant health burden across the globe, causing over 1.6 million deaths annually amongst all ages^1^. The majority of these deaths occur in low- and middle-income countries (LMICs), specifically in south Asia and Sub-Saharan Africa (SSA)^1^. Despite a decrease in global diarrhoeal mortality rates over the past two decades, decreases have not been uniform across age groups or settings^2^. Unsafe water, poor sanitation and hygiene (WaSH)^1^, as well as malnutrition and compromised immunity^3^ are the main risk factors associated with diarrhoeal morbidity and mortality. Other risk factors identified in LMICs include residence in a slum, use of communal toilets and exposure to animals^4^. Children under the age of 5 years are at the highest risk, however there is a significant burden in older age groups, specifically in LMICs^1,5^. Despite global improvements in WaSH^6^, much of the remaining burden of diarrhoeal disease may be attributed to foodborne pathogens^7^. Food safety is of specific concern in LMICs where there is increased consumption of unsafe foods, use of effluent in agriculture and changes in food distribution networks with bulk production and increased distance between production and consumption as well as large, often poorly regulated, informal sectors^7^.

Studies from high-income countries (HICs) have shown the majority of diarrhoeal diseases in adults seeking healthcare to be due to *Campylobacter* spp., *Salmonella* spp., norovirus and rotavirus^8,9^.

Aetiology differs in SSA, with a meta-analysis amongst all ages identifying the main causative pathogens as *Escherichia coli, Cryptosporidium, Cyclospora, Entamoeba* and *Shigella* spp.^10^ There are several reasons for these differences in aetiology including different exposures and risk factors associated with poorer living conditions and poor underlying health. One important consideration is the high prevalence of human immunodeficiency virus (HIV) in SSA^11^. HIV is a known risk factor for diarrhoea, with episodes in HIV-infected individuals more likely to be severe, prolonged and result in hospitalisation with higher mortality rates than in HIV-uninfected individuals^12,13^. The aetiology of diarrhoea in HIV-infected individuals is known to shift with HIV disease progression and treatment^14^.

During early disease stages, while CD4+ counts are high and viral loads low, diarrhoea is often related to HIV seroconversion. As disease progresses, diarrhoea is frequently caused by opportunistic pathogens related to decreased immune function^14^. These pathogens include bacterial infections, such as *Mycobacteria*, parasitic infections, such as *Cystoisospora belli, Cyclospora*, S*trongyloides, Cryptosporidia, Microsporidia* and viral infections, such as cytomegalovirus (CMV)^14–16^. Patients with low CD4+ cell counts are at increased risk for chronic diarrhoea^4^ and polyparasitic infection^17^. The HIV infection itself may be responsible for a proportion of the pathogen-negative cases, through HIV enteropathy and permanent damage to the gastrointestinal mucosa^18^, as well as changes in gut microbial populations leading to dysbiosis^19^. Mucosal damage reduces small bowel villous surface area, causing diarrhoea through malabsorption^20^ as well as defects to cellular and humoral defence mechanisms in the gastrointestinal tract^21^. Pathogen-negative cases associated with advanced HIV disease may also be related to gastrointestinal tract involvement by *Mycobacterium tuberculosis* and/or *avium*^22^ which is difficult to diagnose. A Dutch study found that 34% of diarrhoeal cases in untreated HIV-infected individuals could not be explained by a known causative agent, and hypothesized that these unexplained cases were due to the HIV-infection itself^23^. Initiation of antiretroviral treatment (ART), and accompanying increase in CD4+ cell count, is associated with a shift in the aetiology of diarrhoea from opportunistic infectious causes to non-infectious causes^24^. Data from both HICs and SSA show that although patients initiated on ART have significantly reduced diarrhoeal incidence compared with those not on treatment, the overall diarrhoeal incidence remains high^18,25^. A proportion of the remaining episodes in patients with well-controlled HIV may be drug-related^18^.

There are limited aetiological data on HIV-related diarrhoeal diseases in SSA, with the majority of published studies focusing on children under the age of five years, with available data in older children and adults being limited by diagnostic technology^14^. A review of laboratory records in Botswana indicated that only 14% of stool specimens submitted for routine diagnostic testing for all ages had a pathogen detected, of which 8% were bacteria and 6% parasites^26^. *Shigella* spp. And *Salmonella* spp. were the most commonly detected bacteria while *Cystoisospora* spp. and *Cryptosporidium* spp. were the most commonly detected parasites. Viral testing was not done. Although HIV status was not available for the participants in this analysis, Botswana is a high HIV prevalence setting with an estimated 23.9% of the population between 15-49 years old being HIV-infected during the study time period^26^. Contrary to these findings, a longitudinal cohort study of Zambian adults with a 31% HIV seroprevalence (higher than the estimated population HIV seroprevalence of 22%) found that pathogens could be detected in 99% of stool samples collected (diarrhoeal and non-diarrhoeal patients combined), with *Cryptosporidium* spp., *Cystoisospora* spp. and *Citrobacter* spp. being significantly more common in HIV-infected than HIV-uninfected individuals^5^. They found that HIV-infected adults in the pre-ART era were 2.4 times as likely to suffer from diarrhoea than HIV-uninfected adults and that this increased risk lasted throughout the infection rather than being limited to those with low CD4+ cell counts^5^. Meta-analysis data indicates a high burden of *Cryptosporidium*, microsporidia and *Cystoisospora* in SSA specifically^16^. Another recent meta-analysis identified South Africa as having the highest global *Cryptosporidium* prevalence (57.0%, CI 95%: 24.4-84.5%), although they recognised that these estimates were based on limited data^27^. A study in rural South Africa found that 60% of patients suffering from chronic diarrhoea were HIV-infected and that the majority of these infections were due to *Campylobacter* spp. (20%), *Plesiomonas shigelloides* (17%), *Aeromonas* spp. (13%), *Shigella* spp. (10%), *Salmonella* spp. (10%) and *E. coli* spp. (10%)^28^. Testing did not include parasites or viruses and results were not stratified by ART status. Other studies in African children have identified HIV-infection as a significant risk factor for rotavirus infection^13,29^. Very few studies in SSA in adults included screening for viruses, despite data from HICs showing that viruses (specifically norovirus and rotavirus) were responsible for as much as 44% of diarrhoea in adults presenting to emergency departments^9^. Patients with advanced HIV are at increased risk for diarrhoea due to CMV infection^30^. As CMV is mainly diagnosed on endoscopic biopsy^31^, prevalence is likely underestimated.

South Africa currently has the highest HIV burden in the world, representing 19% of the global HIV-infected population^32^. It is estimated that 86% of HIV-infected adults in South Africa have been diagnosed, with 57% of these on ART (50.8% - 72.7% by province)^33^. Women, elderly patients, those with inadequate access to WaSH facilities and those not yet initiated on ART, have the highest risk for developing diarrhoea amongst HIV-infected South Africans^34^. The majority of data on HIV-related diarrhoea comes from HICs with low HIV prevalence^14^. Since diarrhoeal aetiology is likely to vary between settings^35^, there is a need for more targeted epidemiological studies^18^, specifically in LMICs with high HIV prevalence. There are few studies in HIV-infected populations which investigate a comprehensive range of pathogens or infections with multiple pathogens^19^. These data are important for informing public health interventions and updating diagnostic and treatment guidelines, especially since treatment is often based on guidelines rather than individual patient diagnostics^36^. This study aimed to determine the pathogenic causes for diarrhoeal admissions in HIV-infected patients compared to controls without diarrhoea at three South African hospitals between July 2018 and November 2021.

## Methods

### Case and control enrolment

Patients of all ages hospitalised with diarrhoea were enrolled at Kalafong, Mapulaneng and Matikwana hospitals through the African Network for improved Diagnostics, Epidemiology and Management of common Infectious Agents (ANDEMIA)^37^. Kalafong Provincial Tertiary Hospital is based on the western outskirts of Pretoria (Gauteng Province) and serves the generally low-income communities residing in urban townships to the west of Pretoria^38^. The rural sites included Mapulaneng, a 180 bed district hospital, and Matikwana, a 250 bed regional hospital^39^, both located in Bushbuckridge district in Mpumalanga Province.

Cases were defined as patients admitted with diarrhoea (three or more loose or liquid stools over a 24 hour period) for any duration. Unmatched controls were defined as individuals presenting to the hospital or clinic for reasons other than diarrhoea (vaccination clinic, orthopaedic or surgical wards), without gastrointestinal symptoms (vomiting or diarrhoea) in the past 3 weeks. The study was explained to patients fulfilling the study definition and an information leaflet provided. Written informed consent was signed by patients ≥18 years of age and by parents/guardians for patients <18 years. An assent form was signed for patients between 7-17 years. Surveillance Officers completed investigation forms by interviewing the patient, parent/guardian and reviewing clinical notes and laboratory results where available. HIV status was determined from the clinical notes and available laboratory results. Stool specimens or rectal swabs were collected from both cases and controls.

Patients were enrolled within 48 hours of admission to exclude nosocomial infections. Cases were enrolled from July 2018 to June 2021 and controls enrolled from October 2019 to November 2021. Cases and controls unable to provide specimens and patients <5 years old were excluded from this analysis.

### Laboratory testing

Pathogen screening was performed on Fast-track Diagnostics (viral and bacterial gastroenteritis and stool parasite kits), Seegene Allplex (GI-Parasite and GI-Helminth(I) assays) as well as TaqMan Array Cards (TAC) (included pathogens indicated in Table S1). Monoplex PCR was used to determine final outcome where there were discrepant results between testing platforms for a specific target. Due to limited available TAC cards, cases with unknown HIV status were excluded from testing. Controls with unknown HIV status were included in the overall case-control analysis (since there were limited control specimens available) but were not included in the HIV specific case-control analysis.

### Data management and statistical analysis

Results for CD4+ cell count were extracted from the National Health Laboratory Service (NHLS) database for HIV-infected cases and were categorised into three classes (<200 cells/μl; 200-500 cells/μl; >500 cells/μl). Only CD4+ cell count results within 12 months of the enrolment were included and where multiple results were available for a single patient, the results closest to the time of enrolment were used. Demographic and socioeconomic characteristics for cases and controls were compared. Clinical presentation for HIV-infected versus HIV-uninfected cases were compared. Pathogen detection amongst case and control specimens were compared, stratified by HIV status. Clinical presentation and pathogen detection for HIV-infected cases was compared between CD4+ cell count categories and for ART and cotrimoxazole prophylaxis treatment. X^2^-test and Fisher’s exact test were used for categorical variables and t-test for continuous variables. *P-values*<0.05 were considered to be statistically significant. Stata software (version 14) was used for all analyses.

### Ethical considerations

The ANDEMIA study was approved by the Human Research Ethics Committee (Medical) of the University of the Witwatersrand (approval numbers M170403) and the University of Pretoria Faculty of Health Sciences Research Ethics Committee (approval number 101/2017). The HIV sub-analysis was approved by the Human Research Ethics Committee (Medical) of the University of the Witwatersrand (approval numbers M190663).

## Results

### Enrolment and patient characteristics

A total of 378 specimens for patients ≥5 years were collected during the study period, including 277 cases and 101 controls (Figure 1). Amongst cases, 164 (59.2%) were HIV-infected, 79 (28.5%) were HIV-uninfected and 34 (12.3%) had unknown HIV status. Amongst controls, 23 (22.8%) were HIV-infected, 50 (49.5%) were HIV-uninfected and 28 (27.7%) had unknown HIV status. Cases with unknown HIV status (n=34) were excluded (due to limited testing capacity), hence a total of 344 specimens were included in the analysis.

**Figure 1:**
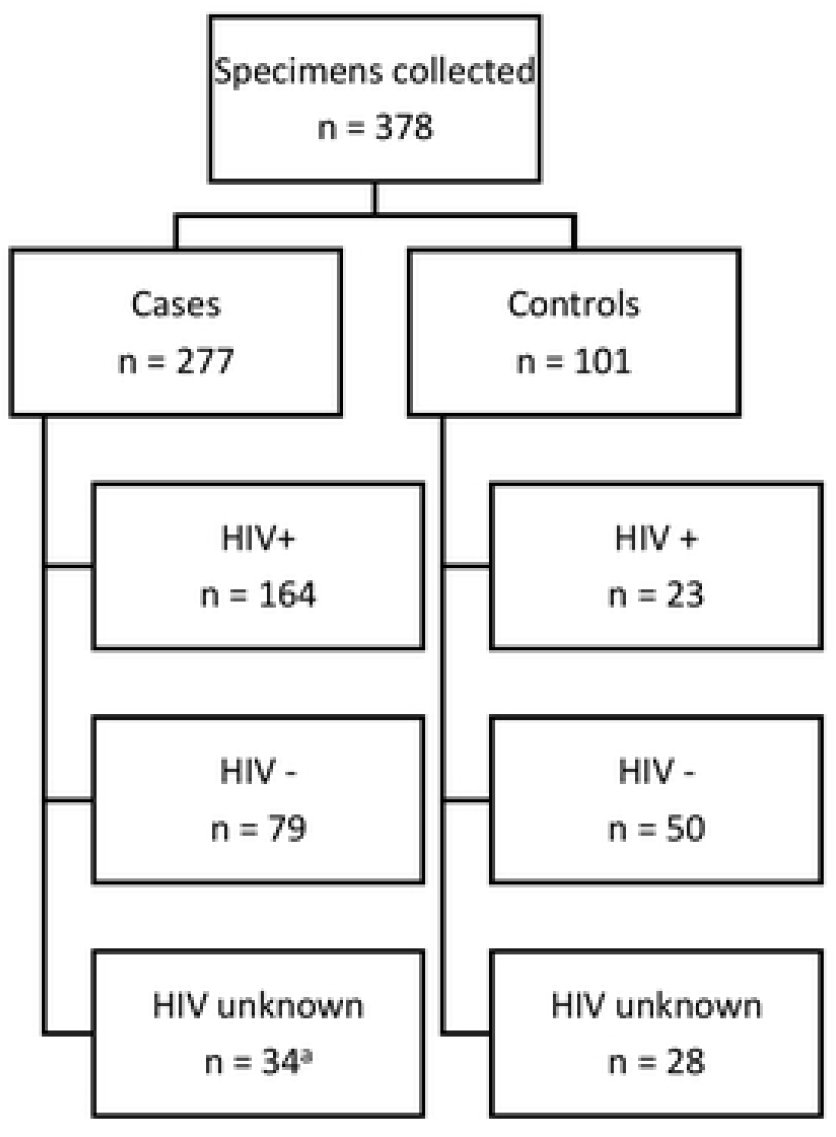
Specimens included in the analysis ^a^Due to limited TAC tests available, cases with unknown HIV status were excluded from the analysis. Controls with unknown HIV status were included to increase sample size for the overall case-control analysis, but were not included in the HIV specific analysis.

The median age of included patients was 34 years (IQR of 23-47), with cases being slightly older (median 36; IQR 26-51) than controls (median 31; IQR 17-38) (Table 1). The majority of patients were female (59.4%) and from the rural sites (67.4%), with proportionally more controls coming from the rural site than cases (82.2% versus 61.3%, *p*<0.001). Cases and controls were similar with respect to household crowding and dietary habits. Both cases and controls had good access to electric or gas cookers (86.1%), refrigerators (88.4%) and improved WaSH (94.8% access to improved water source and 96.4% access to private latrines/flush toilets). CD4+ cell counts were available for 99 (60.4%) of the 164 HIV-infected cases. Eight (8.1%) had counts >500 cells/μl, 21 (21.2%) were between 200-500 cells/μl and the majority (n=70, 70.7%) had <200 cells/μl. Information regarding ART was available for 156 (95.1%) of the HIV-infected cases, of which 130 (83.3%) were on treatment. Mean CD4+ cells counts were similar between those on ART (229.23 cells/μl) and those not on ART (169.7 cells/μl), (*p*=0.418). Information regarding cotrimoxazole treatment was available for 151 HIV-infected patients, of which 63 (41.7%) were on treatment.

**Table 1:**
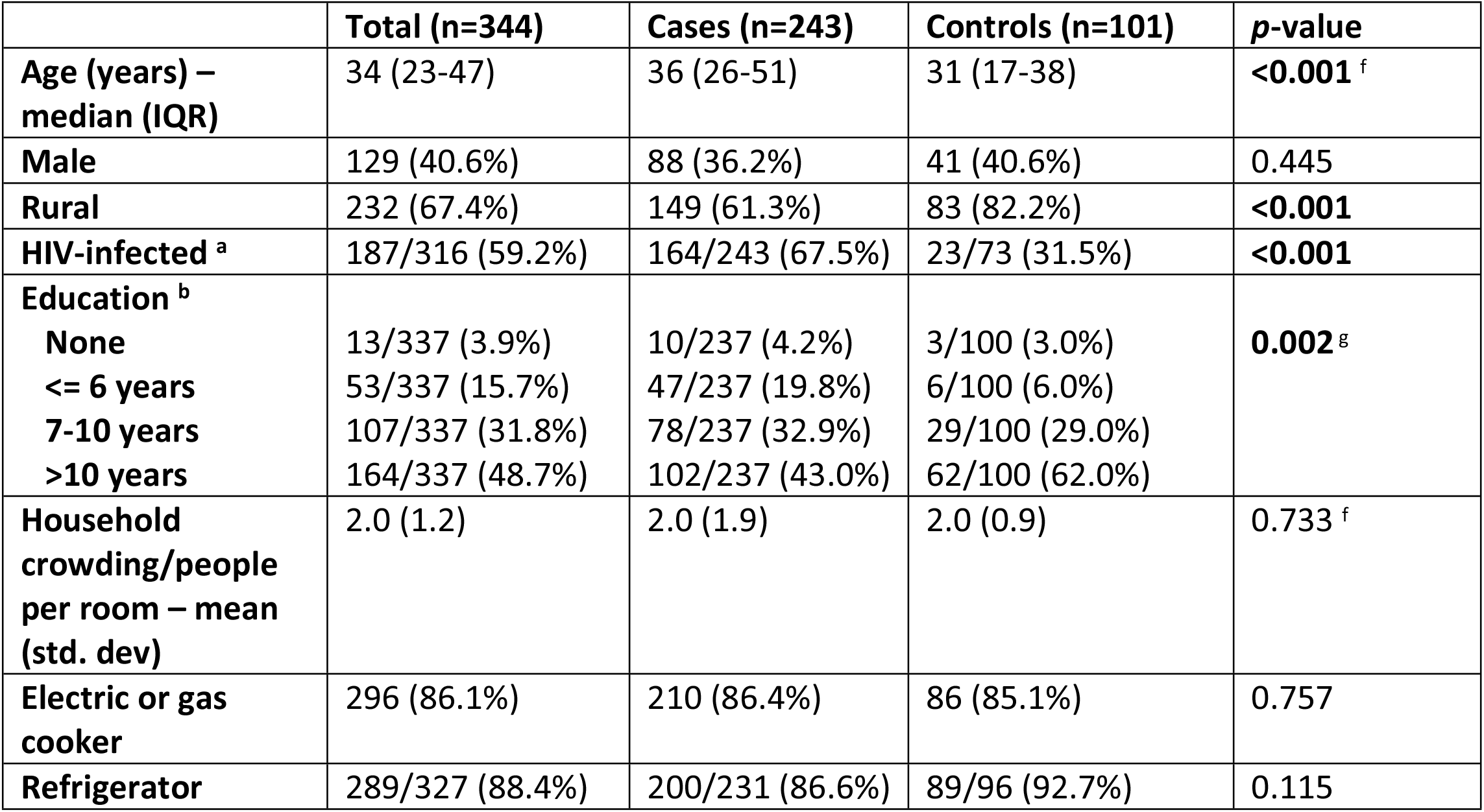
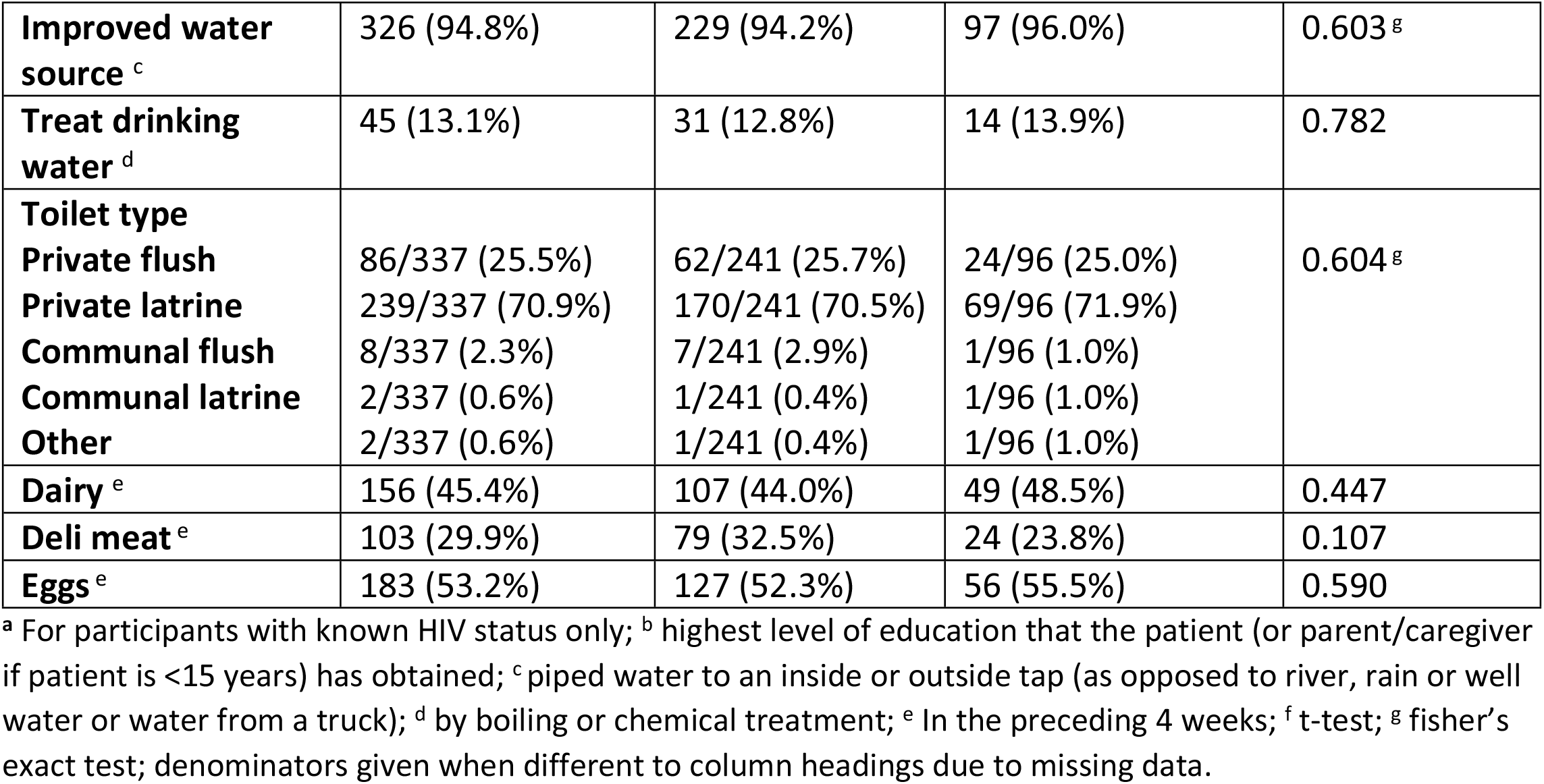
Comparison of patient characteristics for cases and controls

|  | **Total (n=344)** | **Cases (n=243)** | **Controls (n=101)** | **p-value** |
| --- | --- | --- | --- | --- |
| **Age (years) – median (IQR)** | 34 (23-47) | 36 (26-51) | 31 (17-38) | **<0.001<sup>f</sup>** |
| **Male** | 129 (40.6%) | 88 (36.2%) | 41 (40.6%) | 0.445 |
| **Rural** | 232 (67.4%) | 149 (61.3%) | 83 (82.2%) | **<0.001** |
| **HIV-infected<sup>a</sup>** | 187/316 (59.2%) | 164/243 (67.5%) | 23/73 (31.5%) | **<0.001** |
| **Education<sup>b</sup>** |  |  |  | **0.002<sup>g</sup>** |
| **None** | 13/337 (3.9%) | 10/237 (4.2%) | 3/100 (3.0%) |  |
| **<= 6 years** | 53/337 (15.7%) | 47/237 (19.8%) | 6/100 (6.0%) |  |
| **7-10 years** | 107/337 (31.8%) | 78/237 (32.9%) | 29/100 (29.0%) |  |
| **>10 years** | 164/337 (48.7%) | 102/237 (43.0%) | 62/100 (62.0%) |  |
| **Household crowding/people per room – mean (std. dev)** | 2.0 (1.2) | 2.0 (1.9) | 2.0 (0.9) | 0.733<sup>f</sup> |
| **Electric or gas cooker** | 296 (86.1%) | 210 (86.4%) | 86 (85.1%) | 0.757 |
| **Refrigerator** | 289/327 (88.4%) | 200/231 (86.6%) | 89/96 (92.7%) | 0.115 |
| **Improved water source<sup>c</sup>** | 326 (94.8%) | 229 (94.2%) | 97 (96.0%) | 0.603<sup>g</sup> |
| **Treat drinking water<sup>d</sup>** | 45 (13.1%) | 31 (12.8%) | 14 (13.9%) | 0.782 |
| **Toilet type** |  |  |  |  |
| Private flush | 86/337 (25.5%) | 62/241 (25.7%) | 24/96 (25.0%) | 0.604<sup>g</sup> |
| Private latrine | 239/337 (70.9%) | 170/241 (70.5%) | 69/96 (71.9%) |  |
| Communal flush | 8/337 (2.3%) | 7/241 (2.9%) | 1/96 (1.0%) |  |
| Communal latrine | 2/337 (0.6%) | 1/241 (0.4%) | 1/96 (1.0%) |  |
| Other | 2/337 (0.6%) | 1/241 (0.4%) | 1/96 (1.0%) |  |
| **Dairy<sup>e</sup>** | 156 (45.4%) | 107 (44.0%) | 49 (48.5%) | 0.447 |
| **Deli meat<sup>e</sup>** | 103 (29.9%) | 79 (32.5%) | 24 (23.8%) | 0.107 |
| **Eggs<sup>e</sup>** | 183 (53.2%) | 127 (52.3%) | 56 (55.5%) | 0.590 |

<sup>a</sup> For participants with known HIV status only; <sup>b</sup> highest level of education that the patient (or parent/caregiver if patient is <15 years) has obtained; <sup>c</sup> piped water to an inside or outside tap (as opposed to river, rain or well water or water from a truck); <sup>d</sup> by boiling or chemical treatment; <sup>e</sup> In the preceding 4 weeks; <sup>f</sup> t-test; <sup>g</sup> fisher's exact test; denominators given when different to column headings due to missing data.

### Clinical presentation of cases

HIV-infected cases were likely to experience symptoms for a longer duration before admission (median of 5 days versus 2 days, *p*<0.001) and were more likely to have chronic or persistent diarrhoea (8.5% versus 0.0%, *p*=0.006) than HIV-uninfected cases (Table 2). The most common symptoms experienced by all cases were fatigue (182, 74.9%), weight loss (176, 72.4%), vomiting (172, 70.8%), fever (168, 69.1%), abdominal pain (166, 68.3%) and nausea (165, 67.9%). HIV-infected cases were more likely to suffer from weight loss (81.1% versus 54.4%, p<0.001) and nausea (72.0% versus 59.5%, *p*=0.051), while HIV-uninfected cases were more likely to suffer from dysentery (15.2% versus 4.3%, *p*=0.003).

**Table 2:** Clinical presentation of diarrhoeal cases

|  | **Total (n=243)** | **HIV-infected (n=164)** | **HIV-uninfected (n=79)** | **p-value** |
| --- | --- | --- | --- | --- |
| **Duration of symptoms before admission (days) – median (IQR)** | 4 (2-8) | 5 (3-8) | 2 (1-4) | **<0.001<sup>d</sup>** |
| **Chronic/persistent diarrhoea<sup>a</sup>** | 13 (5.4%) | 13 (7.9%) | 0 (0.0%) | **0.011<sup>e</sup>** |
| **Fatigue** | 182 (74.9%) | 124 (75.6%) | 58 (73.4%) | 0.712 |
| **Weight loss** | 176 (72.4%) | 133 (81.1%) | 43 (54.4%) | **<0.001** |
| **Vomiting** | 172 (70.8%) | 118 (72.0%) | 54 (68.4%) | 0.564 |
| **Fever<sup>b</sup>** | 168 (69.1%) | 113 (68.9%) | 55 (69.6%) | 0.910 |
| **Abdominal pain** | 166 (68.3%) | 106 (64.6%) | 60 (76.0%) | 0.076 |
| **Nausea** | 165 (67.9%) | 118 (72.0%) | 47 (59.5%) | 0.051 |
| **Respiratory symptoms** | 105 (43.2%) | 77 (47.0%) | 28 (35.4%) | 0.090 |
| **Headache** | 100 (41.2%) | 68 (41.5%) | 32 (40.5%) | 0.887 |
| **Chills** | 63 (25.9%) | 48 (29.3%) | 15 (19.0%) | 0.087 |
| **Arthralgia** | 51 (21.0%) | 38 (23.2%) | 13 (16.5%) | 0.229 |
| **Dermatological symptoms** | 31 (12.8%) | 25 (15.2%) | 6 (7.6%) | 0.094 |
| **Myalgia** | 24 (10.0%) | 16 (9.8%) | 8 (10.1%) | 0.928 |
| **Dysentery <sup>c</sup>** | 19 (7.8%) | 7 (4.3%) | 12 (15.2%) | **0.003** |
| **Neurological symptoms** | 10 (4.1%) | 6 (3.7%) | 4 (5.1%) | 0.732 <sup>e</sup> |
| **Painful lymphadenopathy** | 4 (1.7%) | 3 (1.9%) | 1 (1.3%) | >0.99 <sup>e</sup> |

<sup>a</sup> diarrhoea for 28 days or longer; <sup>b</sup> current temperature of >38°C or history of fever in the past 10 days; <sup>c</sup>
Dysentery defined as self-reported blood in the stool; <sup>d</sup> t-test; <sup>e</sup> fisher's exact test

### Pathogen detection in specimens of cases and controls

Pathogens were detected in 66.3% (228) of specimens tested, with significantly higher detection in cases compared with controls (72.8% versus 50.5%, *p*<0.001; Table 3). Bacteria were more prevalent in cases compared with controls (50.2% versus 28.7%, *p*<0.001), specifically *Shigella* spp., *Salmonella* spp. and *Helicobacter pylori* (24.3% versus 10.9%, *p*=0.005; 8.6% versus 1.0%, *p*=0.007 and 8.6% versus 0.0%, *p*=0.001, respectively). Detection of viruses was significantly higher in cases compared with controls (21.8% versus 10.9%, *p*=0.018), specifically for adenovirus (11.0% versus 4.0%, *p*=0.038). Parasites were detected in 37.8% (130) of specimens tested, however, detection was not significantly higher in cases than controls (40.7% versus 30.7%, *p*=0.080) with the available sample size. Prevalence of *Cystoisospora, Cryptosporidium* spp. and *Enterocytozoon* spp. was significantly higher in cases than controls (11.9% versus 0.0%, *p*<0.00; 11.1% versus 3.0%, *p*=0.012; 4.5% versus 0.0%, *p*=0.038, respectively). *Blastocystis* and *Schistosoma* were more prevalent amongst controls than amongst cases (23.8% versus 10.3%, *p*=0.001; 6.9% versus 2.1%, *p*=0.025, respectively). Cases were more likely than controls to have multiple pathogens detected (37.9% versus 22.8%, *p*=0.007). Amongst HIV-infected participants, *Cystoisospora* spp. prevalence was significantly higher in cases than in controls (17.7% versus 0.0%, *p*=0.028), while *Schistosoma* was detected more often in controls compared with cases (17.4% versus 2.4%, *p*=0.009). Amongst the HIV-uninfected participants, *Shigella* spp., *Salmonella* spp. and *Helicobacter pylori* were more prevalent amongst cases than controls (36.7% versus 12.0%, *p*=0.002; 11.4% versus 0.0%, *p*=0.012; 10.1% versus 0.0%, *p*=*0*.023). Norovirus GII, *C. difficile, Campylobacter, Cystoisospora, Cryptosporidium* spp. and *Enterocytozoon* spp. were more prevalent amongst HIV-infected than HIV-uninfected participants (6.4% versus 1.6%, *p*=0.032; 7.0% versus 0.8%, *p*= 0.010; 6.4% versus 0.8%, *p*= 0.018; 15.5% versus 0.01%, *p*<0.001; 15.0% versus 1.6%, *p*<0.001; 5.4% versus 0.8%, *p*=0.031). *Shigella* spp. and *Blastocystis* were more prevalent amongst HIV-uninfected than HIV-infected participants (27.1% versus 17.1%, *p*=0.032; 25.6% versus 8.0%, *p*<0.001). *Shigella* spp. (18.3%) and *Cystoisospora* (17.7%) were the most prevalent pathogens detected in HIV-infected diarrhoeal cases, while *Shigella* spp. (36.7%) and *Salmonella* spp. (11.4%) were the most prevalent pathogens in HIV-uninfected diarrhoeal cases. Although *Blastocystis* was prevalent in HIV-uninfected cases, it was not associated with diarrhoea as prevalence was significantly higher in HIV-uninfected controls (40.0% versus 16.5%, *p*<0.001).

**Table 3:**
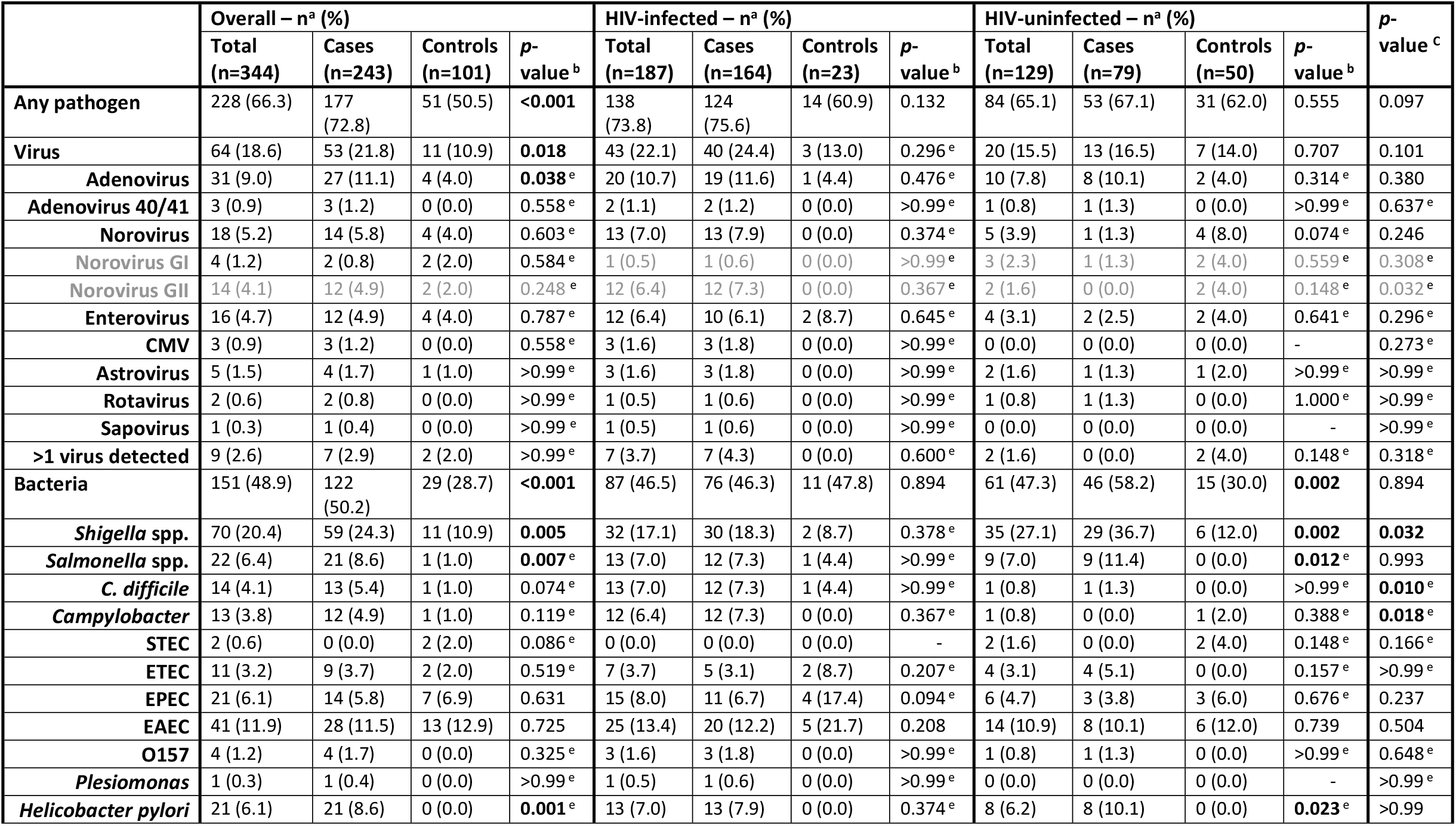
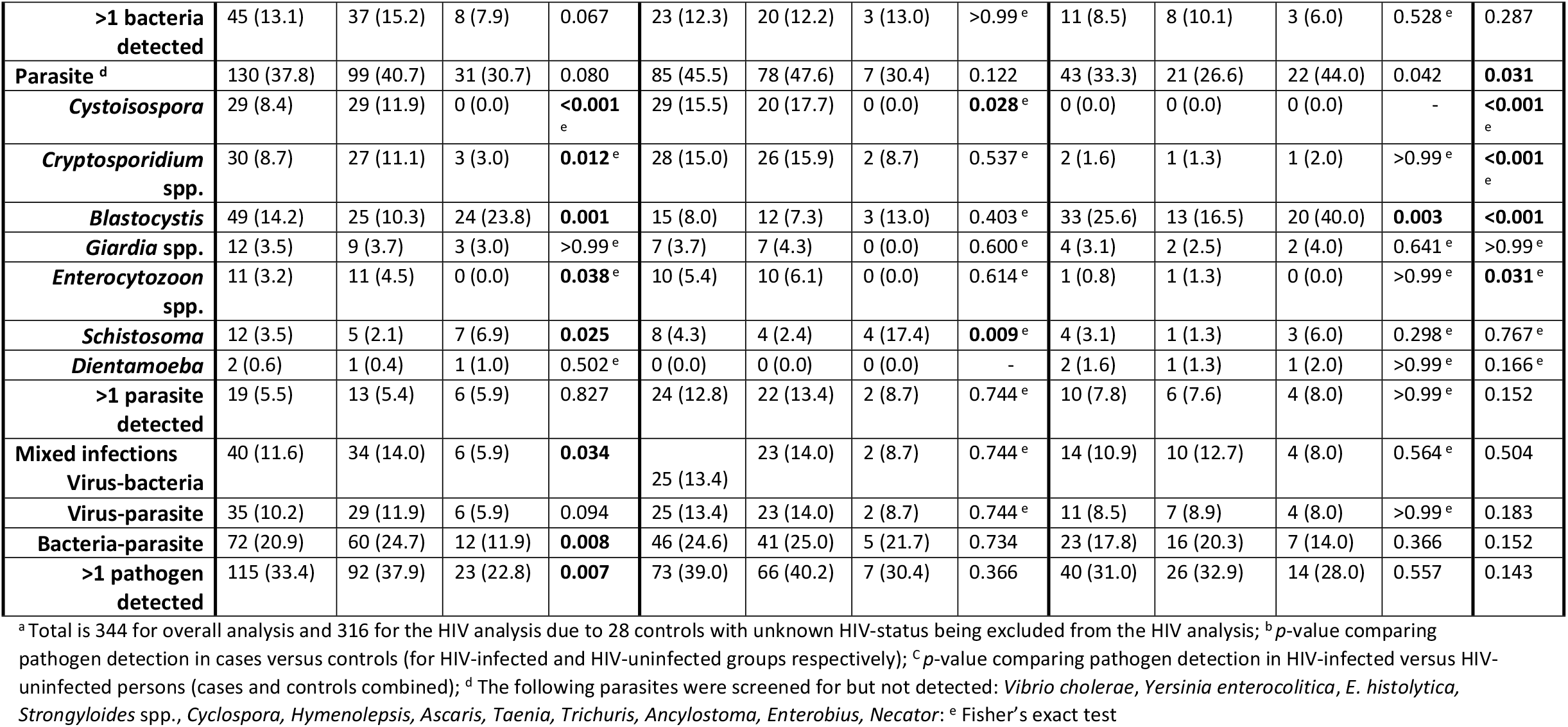
Pathogen detection in specimens of cases and controls, stratified by HIV-infection status

### Effect of CD4+ cell count and treatment on clinical presentation and pathogen detection amongst HIV-infected diarrhoeal cases

For HIV-infected cases, presentation did not differ by CD4+ cell count for duration of symptoms, fever, vomiting, fatigue, nausea, respiratory symptoms, abdominal pain, dermatological symptoms, chills, headache, dysentery, arthralgia, myalgia, neurological symptoms or painful lymphadenopathy (Table S2). There was an increased proportion of cases reporting weight loss with lower CD4+ cell counts (91.4%, 81.0% and 62.5% for CD4+ cell counts of <200, 200-500, >500 cells/mm^3^ respectively; *p*=0.032). Pathogens were detected more frequently in cases with low CD4+ cell counts (75.7%, 42.9% and 37.5% for CD4+ cell counts of <200, 200-500, >500 cells/mm^3^ respectively; *p*=0.004) (Table S3). *Cryptosporidium* spp. specifically, was detected more often in those with low CD4+ cell counts (25.7%, 4.8% and 0.0% for CD4+ cell counts of <200, 200-500, >500 cells/mm^3^ respectively; *p*=0.039).

Clinical presentation was similar amongst HIV-infected cases on ART and those not on ART (Table S4). Those on cotrimoxazole prophylaxis were more likely to experience fatigue (88.9% versus 69.3%, *p*=0.005), weight loss (90.5% versus 76.1%, *p*=0.023) and chills (50.8% versus 15.9%, *p*<0.001) then those not on cotrimoxazole. Pathogen detection was similar amongst HIV-infected cases on ART and those not on ART (72.3% versus 88.5%, *p*=0.082) (Table S5). Parasites, specifically *Cystoisospora* spp. were more commonly detected amongst patients on cotrimoxazole compared with those not on cotrimoxazole (27.0% versus 11.4%, *p*=0.014). Other pathogens were found in similar proportions across treatment groups.

### Clostridioides difficile detection

*C. difficile* was detected in 14 cases, 13 of which were in HIV-infected patients (92.9%). Symptoms were similar between cases in which *C. difficile* was detected and those without *C. difficile* detection, however those with *C. difficile* detected were more likely to have had a longer duration of illness before admission (6.5 (IQR 4-11) days versus 4 (IQR 2-7) days) (data not shown). It is important to distinguish *C. difficile* colonisation from disease to avoid unnecessary antimicrobial therapy. The case definition, as per the guidelines for diagnostic and clinical management of *C. difficile* from the South African Society for Clinical Microbiology, includes onset of diarrhoea more than 48 hours after admission, or diarrhoea that continues for 3 days post admission and where there is no likely alternatively cause (no other pathogens detected or use of laxatives), or where the patient has had an admission or antibiotics use in the past 12 weeks.^40^ Three of the 14 cases did not match this case definition as they did not have an admission in the past 12 weeks and had multiple pathogens detected on PCR. These three cases were however HIV-infected. Five of the cases had no other pathogens detected on PCR, all of which were HIV-infected. Eight of the cases did have a prior admission, however the majority of these (n=6, 75.0%) had multiple pathogens detected on PCR. In addition, the guidelines recommend that specimens testing positive for *C. difficile* on PCR (which is highly sensitive for detection of C. *difficile*), should have further toxins A/B immunoassays performed (which have high specificity for diagnosis of *C. difficile* infection). This was not done in the current analysis.

## Discussion

Despite advances in HIV treatment, diarrhoea amongst HIV-infected patients remains a significant public health challenge^19^. Our study found that the majority (67.5%) of patients 5 years and older with known HIV-status admitted to the study sites with diarrhoea were HIV-infected, highlighting the importance of HIV-related illnesses among South African adults hospitalised with diarrhoea. The Joint United Nations Programme on HIV/AIDS (UNAIDS) 90-90-90 treatment target aims to ensure that 90% of the HIV-infected population are diagnosed, 90% of those diagnosed are initiated on ART and 90% of those on ART should achieve viral suppression^41^. A recent South African analysis estimated that 70.7% of diagnosed HIV-infected individuals have initiated ART, and 87.4% of these are thought to be virally suppressed^41^. Proportions reported in the current analysis reflect these numbers, as 83.3% of HIV-infected patients with known ART status were on treatment, however a large proportion of HIV-infected patients included in this analysis had CD4+ cell count <200 cells/μl (70/99, 70.7%) indicating that they are unlikely to be virally suppressed. Mpumalanga (home province to the majority of the patients included in this analysis) is the only South African province in which the third-90 indicator value (the percentage of those on ART that are virally suppressed) is below 85%^41^. It is also likely that the study design was skewed towards detection of poorly suppressed HIV cases as they are most likely to be hospitalised for diarrhoea. Diarrhoea in patients with viral suppression will likely be seen at a clinic level. Further studies at clinic level are recommended in order to include a broader spectrum of cases. The clinical presentation of diarrhoea in HIV-infected cases differs to that of HIV-uninfected cases. HIV-infected diarrhoeal cases were more likely to have symptoms for a longer duration before admission and to suffer from weight loss and nausea. HIV-uninfected cases were more likely to suffer from dysentery. This finding has not been reported in other studies, however it is likely related to aetiological differences between HIV-infected and HIV-uninfected diarrhoeal patients. Amongst HIV-infected cases, patients on cotrimoxazole were more likely to experience fatigue, weight loss and chills and were more likely to have parasites detected, specifically *Cystoisospora* spp., compared to those not on cotrimoxazole. This is an interesting finding since cotrimoxazole is the prescribed treatment for this pathogen. Since all patients with CD4+ <200 cells/μl should receive cotrimoxazole prophylaxis, it can be used as a surrogate marker for advanced HIV disease. However this finding may also point to changes in *Cystoisospora* sensitivity for cotrimoxazole or may indicate that these patients are carriers or take longer to clear the pathogen. The possibility of drug resistance should be further investigated in future studies.

The use of molecular methods and the expanded panel of pathogens included in testing resulted in a high pathogen detection rate, with pathogen detection being significantly higher in cases compared with controls. Although viruses were commonly detected (18.5% of all specimens tested) and were more likely to be detected in cases as compared with controls, when stratified by HIV status, viruses were not significantly associated with diarrhoea in this study. Although this lack of association may be partly due to asymptomatic carriage of viruses (specifically in HIV-uninfected patients), it is likely that the sample size was insufficient to determine significant differences between cases and controls when stratified by HIV status. Interestingly, norovirus GII was more prevalent in HIV-infected individuals than HIV-uninfected individuals and detection in HIV-infected individuals was limited to cases. Previous findings from South Africa have shown that HIV-infected children are more likely to suffer poor outcomes related to norovirus infection^42^, however detection rates were similar amongst HIV-infected and –uninfected children. This increased detection of norovirus in HIV-infected patients ≥5 years of age has not been previously described. Bacteria was detected in 43.9% of specimens tested and was significantly associated with diarrhoea overall and amongst HIV-uninfected patients. Interestingly, prevalence of bacterial pathogens was high in both cases and controls within the HIV-infected group. This was specifically due to the high prevalence of *E. coli* spp. (including ETEC, EPEC and EAEC) amongst HIV-infected controls. High *E. coli* spp. colonisation rates in HIV-infected patients have been documented in other settings^43^. We found *Shigella* spp, *Salmonella* spp. and *H. pylori* to be significantly associated with diarrhoea in the current study, specifically amongst HIV-uninfected patients. It’s likely that the sample size was insufficient to detect a difference in prevalence between cases and controls in the HIV-uninfected group. *Campylobacter* spp. and *C. difficile* were detected more often in HIV-infected patients as compared with HIV-uninfected patients but were not significantly associated with diarrhoea (likely due to small sample sizes).

As expected from literature^14^, parasite prevalence was significantly higher amongst HIV-infected compared with HIV-uninfected patients, specifically *Cystoisospora, Cryptosporidium* spp. and *Enterocytozoon* spp., although only *Cystoisospora* was significantly associated with diarrhoea amongst HIV-infected patients. The case-control analysis indicated that *Blastocystis* and *Schistosoma* are unlikely to be causative agents of diarrhoea since these pathogens were detected at higher rates in controls than in cases. Prevalence of *Schistosoma* among healthy individuals has been reported to be as high as 51.2% in the Democratic Republic of Congo and is likely to be higher in villages and rural areas where water is collected from rivers and dam^44^. *Blastocystis* has been noted as an important cause of diarrhoea amongst immunosuppressed patients^45^, however this was not found in the current study.

Diarrhoeal aetiology differed with HIV status, with *Shigella* spp. and *Salmonella* spp. being prevalent amongst HIV-uninfected cases and *Shigella* spp., *Cystoisospora*, and *Cryptosporidium* spp. being prevalent amongst HIV-infected cases. These differences should be considered during development of diagnostic and treatment guidelines. This study highlights the importance of *Shigella* spp. in diarrhoeal morbidity amongst adults (regardless of HIV status). According to the Global Burden of Disease Study (GBD), *Shigella* spp. were the leading cause of diarrhoeal deaths amongst individuals over the age of 5 years and the second leading cause of death in young children in 2016^1^. While there is currently no approved *Shigella* vaccine, there are several candidates which show promise for efficacy testing^46^. In order to introduce a candidate vaccine, burden estimates are required. Although a larger sample size is required, this analysis gives an indication of the high burden of *Shigella* spp. in our setting. Further work is required to determine if HIV-infected individuals should be considered as a risk group for possible target should *Shigella* vaccine become available.

Our results are in line with clinical literature which indicates a shift in diarrhoeal aetiology with CD4+ cell count amongst HIV-infected patients. Pathogen detection was high in those with CD4+ cell counts <200 cells/μl, indicating increased opportunistic infections (specifically *Cryptosporidium* spp.). Pathogens could only be detected in just over a third of cases with high CD4+ cell count (>500 cells/μl), suggesting that these cases are possibly drug-related. This finding may be useful for treatment guidelines. It also highlights the importance of addressing diarrhoea in patients on ART especially since chronic diarrhoea has been identified as an major cause of dropout from ART care services^47^.

Detection of *C. difficile* in this study was almost exclusive to HIV-infected participants (13/14, 92.9%; including 12 cases and 1 control). Data from another hospital study in Gauteng Province estimated that community acquired *C. difficile* infections represent only 1.3% of cases (with the remaining 98.7% being hospital acquired)^48^. Our study design excluded cases of hospital-acquired diarrhoea, as enrolment and specimen collection was done within 48 hours of admission. The 5.4% *C. difficile* prevalence reported here is lower than systematic review estimates of 15.8% amongst symptomatic non-immunosuppressed in-patients from LMIC^49^. This systematic review indicated similar rates amongst immunosuppressed and non-immunosuppressed populations^49^ whereas the current study found that *C. difficile* was more commonly detected in HIV-infected cases than in –uninfected cases. While not considered an opportunistic infection, *C. difficile* is associated with exposure to healthcare settings and with the use of antibiotics, both of which are increased amongst HIV-patients^50^. The increased management of HIV patients in an outpatient setting^51^ may dispute the inclusion of recent admission as a criterion in the case definition. There is also growing evidence to suggest the possibility of *C. difficile* being an important community-acquired pathogen^51^, and consideration of this should be given in populations with high HIV prevalence.

A major limitation of this study was the small sample size which restricted the analysis (attributable fractions and odds ratios could not be calculated as many categories had zero count). The ANDEMIA study was not specifically aimed at enrolling HIV-infected cases and controls, therefore this analysis relied on incidental HIV enrolments. Enrolment, specifically of controls, was also hampered by COVID-19 restrictions. An extension of this analysis would benefit from enrolling controls from HIV clinics. Viral load was not analysed here, since few recent results were available; thus recent CD4+ cell count was used as a proxy. CD4+ cell counts were not available for all HIV-infected cases at the time of admission. Many of those without CD4+ cell counts available were matched from NHLS data, however these tests were not always performed at the same time as the enrolment. We used a cut off of 12 months on either side of enrolment for CD4+ cell counts as this was deemed to be a reasonable estimation of the CD4+ cell count at admission. In cases where there were significant changes in CD4+ cell counts over time, these may have been incorrectly quantified. There were also limited data available for antibiotic use. Date of last dose was available, however start date for antibiotic course was not collected. This has limitations specifically for the *C. difficile* analysis, since we could not determine the length of antibiotic courses. Future studies should also investigate the expansion of diagnostic testing for *M. avium* which was not done in the present study.

In conclusion, this study highlights the importance of HIV-related diarrhoea amongst South African inpatients ≥5 years of age, and underscores the importance of research in a high HIV prevalence setting in order to improve the understanding of aetiology of diarrhoea. These data are important for development of guidelines and treatment protocols as well as for preventative interventions, such as vaccine introduction. Our data specifically highlights the importance of *Shigella* spp. amongst both HIV-infected and HIV-uninfected diarrhoeal patients. We suggest that research be expanded to primary healthcare levels in order to include a broader spectrum of HIV disease, specifically to investigate diarrhoea in individuals with well-controlled HIV.

## Data Availability

The datasets generated and/or analysed during the current study are not publicly available as they include study data which has not yet been published. Data are available from the corresponding author on reasonable request.

## Acknowledgments

We wish to acknowledge the ANDEMIA participants, funders, staff and clinical staff at the hospitals. Thank you to Ann Christin Vietor for statistical assistance.

## Supplementary tables

**Table S1:** Pathogens included in molecular testing

| Pathogen | Testing platform |
| --- | --- |
| **Viruses** |  |
| Norovirus GI | FTD viral gastroenteritis, TaqMan Array Cards |
| Norovirus GII | FTD viral gastroenteritis, TaqMan Array Cards |
| Human astrovirus | FTD viral gastroenteritis, TaqMan Array Cards |
| Rotavirus | FTD viral gastroenteritis, TaqMan Array Cards |
| Human adenovirus | FTD viral gastroenteritis, TaqMan Array Cards |
| Sapovirus | FTD viral gastroenteritis, TaqMan Array Cards |
| Enterovirus | TaqMan Array Cards |
| CMV | Monoplex PCR |
| **Bacteria** |  |
| *Campylobacter coli/jejuni/lari* | FTD bacterial gastroenteritis, TaqMan Array Cards |
| *Clostridioides difficile* | FTD bacterial gastroenteritis, TaqMan Array Cards |
| *Escherichia coli* verotoxin positives | FTD bacterial gastroenteritis |
| *Salmonella* spp. | FTD bacterial gastroenteritis, TaqMan Array Cards |
| *Shigella* spp. | FTD bacterial gastroenteritis, TaqMan Array Cards |
| Enteroinvasive *Escherichia coli* | FTD bacterial gastroenteritis, TaqMan Array Cards |
| *Yersinia enterocolitica* | FTD bacterial gastroenteritis, TaqMan Array Cards |
| EAEC | TaqMan Array Cards |
| EPEC | TaqMan Array Cards |
| ETEC | TaqMan Array Cards |
| STEC/VTEC | FTD bacterial gastroenteritis; TaqMan Array Cards |
| *E. coli* O157 | TaqMan Array Cards |
| **Parasites** |  |
| *Plesiomonas shigelloides* | TaqMan Array Cards |
| *Entamoeba histolytica* | FTD enteric parasites, TaqMan Array Cards, SeeGene Allplex |
| *Cryptosporidium* spp. | FTD enteric parasites, TaqMan Array Cards, SeeGene Allplex |
| *Giardia lamblia* | FTD enteric parasites, TaqMan Array Cards, SeeGene Allplex |
| *Vibrio cholerae* | TaqMan Array Cards |
| *Helicobacter pylori* | TaqMan Array Cards |
| *Schistosoma* | TaqMan Array Cards |
| *Enterocytozoon* spp. | TaqMan Array Cards, SeeGene Allplex |
| *Strongyloides stercoralis* | TaqMan Array Cards, SeeGene Allplex |
| *Cytoisospora belli* | TaqMan Array Cards |
| *Blastocystis hominis* | SeeGene Allplex |
| *Dientamoeba fragilis* | SeeGene Allplex |
| *Cyclospora cayentanensis* | SeeGene Allplex |
| *Hymenolepis nana* | SeeGene Allplex |
| *Ascaris lumbricoides* | SeeGene Allplex |
| *Taenia* spp. | SeeGene Allplex |
| *Trichuris trichiura* | SeeGene Allplex |
| *Ancylostoma duodenale* | SeeGene Allplex |
| *Enterobius vermicularis* | SeeGene Allplex |
| *Necator americanus* | SeeGene Allplex |

**Table S2:** Clinical presentation for HIV-infected cases, stratified by CD4+ cell counts

|  | CD4+ cell count <sup>a</sup> |  |  | *p*-value |
| --- | --- | --- | --- | --- |
|  | <200 cells/μl<br>(n=70) | 200-500 cells/μl<br>(n=21) | >500 cells/μl<br>(n=8) |  |
| Duration of symptoms before admission – median (IQR) | 6 (3-10) | 7 (3.5-12) | 5.5 (3-7) | 0.822 |
| Chronic/persistent diarrhoea | 6 (8.6%) | 2 (9.5%) | 1 (12.5%) | 0.855 |
| Weight loss | 64 (91.4%) | 17 (81.0%) | 5 (62.5%) | **0.032** |
| Fatigue | 58 (82.9%) | 16 (76.2%) | 5 (62.5%) | 0.293 |
| Nausea | 6 (75.0%) | 15 (71.4%) | 6 (75.0%) | >0.99 |
| Vomiting | 48 (68.6%) | 14 (66.7%) | 6 (75.0%) | >0.99 |
| Fever (current or history) | 46 (65.7%) | 14 (66.7%) | 5 (62.5%) | >0.99 |
| Abdominal pain | 41 (58.6%) | 14 (66.7%) | 4 (50.0%) | 0.667 |
| Respiratory symptoms | 33 (47.1%) | 12 (57.1%) | 1 (12.5%) | 0.097 |
| Chills | 32 (45.7%) | 7 (33.3%) | 3 (37.5%) | 0.634 |
| Headache | 32 (45.7%) | 7 (33.3%) | 4 (50.0%) | 0.635 |
| Dermatological symptoms | 13 (18.6%) | 3 (14.3%) | 0 (0.0%) | 0.494 |
| Arthralgia | 11 (15.7%) | 6 (28.6%) | 0 (0.0%) | 0.171 |
| Myalgia | 11 (15.7%) | 1 (4.8%) | 0 (0.0%) | 0.334 |
| Neurological symptoms | 5 (7.1%) | 0 (0.0%) | 1 (12.5%) | 0.347 |
| Dysentery <sup>b</sup> | 2 (2.9%) | 0 (0.0%) | 0 (0.0%) | >0.99 |
| Painful swollen glands | 2 (2.9%) | 0 (0.0%) | 0 (0.0%) | >0.99 |

<sup>a</sup> CD4+ count within 12 months of enrolment only known for 99 of the 164 HIV-infected patients included in molecular testing; <sup>b</sup> Dysentery defined as self-reported blood in the stool.

**Table S3:**
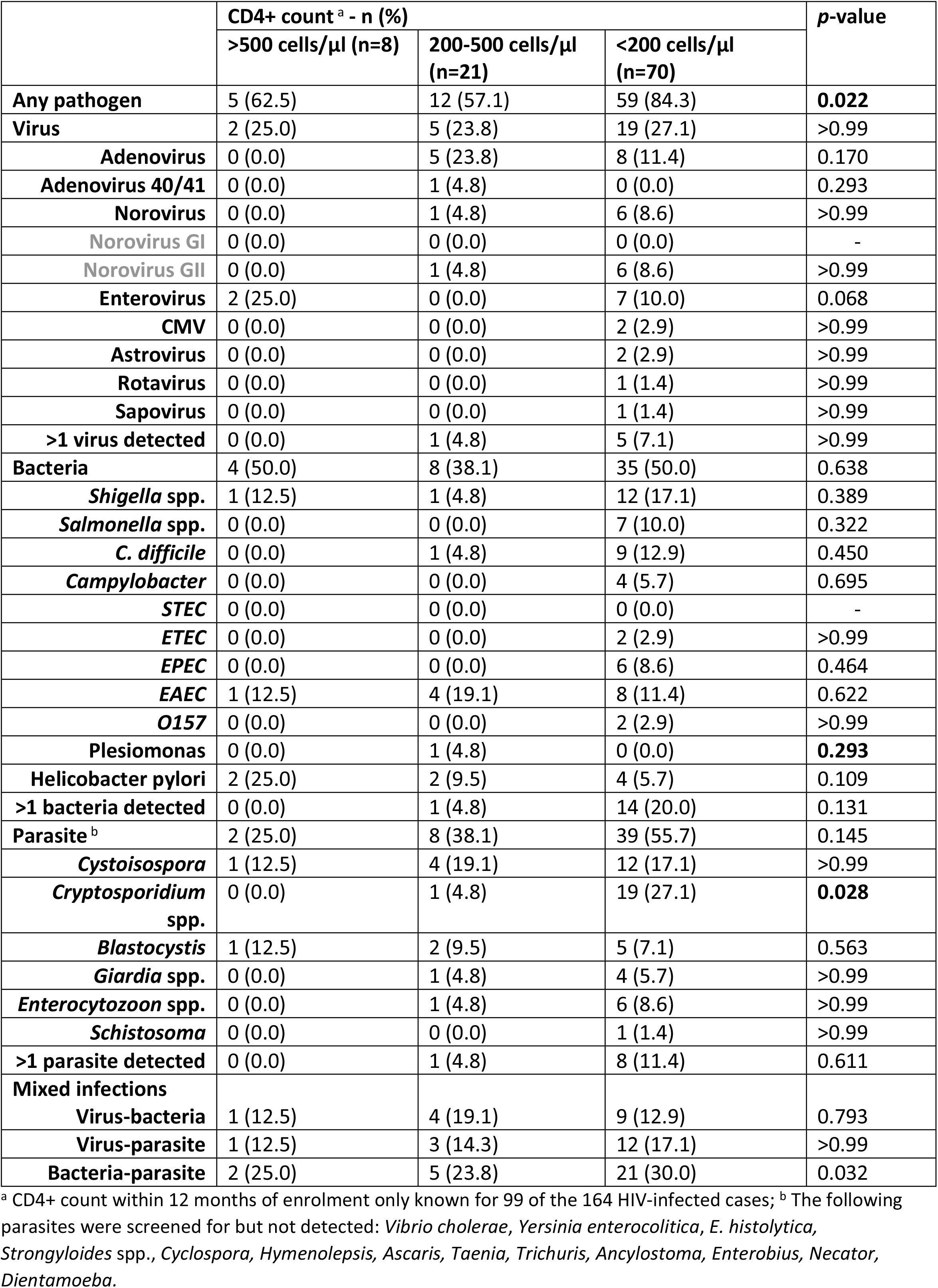
Pathogen detection in specimens of HIV-infected cases, stratified by CD4+ cell count

**Table S4:**
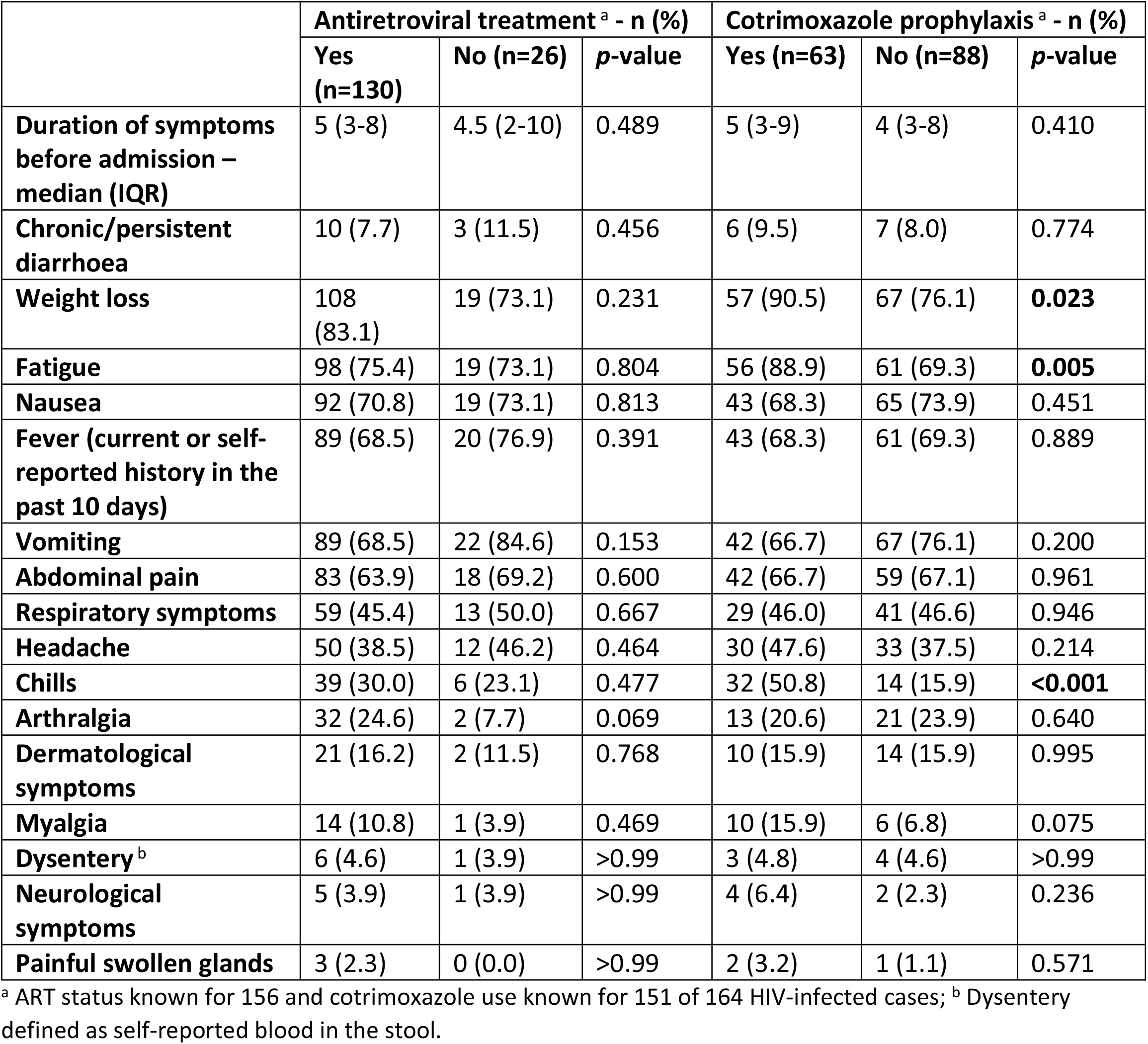
Clinical presentation of HIV-infected cases, stratified by treatment

**Table S5:** Pathogen detection in specimens of HIV-infected cases, stratified by treatment

|  | Antiretroviral treatment<sup>a</sup> - n (%) |  |  | Cotrimoxazole prophylaxis<sup>a</sup> - n (%) |  |  |
| --- | --- | --- | --- | --- | --- | --- |
|  | No (n=26) | Yes (n=130) | p-value | No (n=88) | Yes (n=63) | p-value |
| **Any pathogen** | 23 (88.5) | 94 (72.3) | 0.082 | 66 (75.0) | 49 (77.8) | 0.693 |
| **Virus** | 6 (23.1) | 32 (24.6) | 0.868 | 20 (22.7) | 16 (25.4) | 0.704 |
| **Adenovirus** | 3 (11.5) | 14 (10.8) | >0.99 | 9 (10.2) | 8 (12.7) | 0.795 |
| **Adenovirus 40/41** | 0 (0.0) | 1 (0.8) | >0.99 | 1 (1.1) | 0 (0.0) | >0.99 |
| **Norovirus** | 3 (11.5) | 10 (7.7) | 0.456 | 6 (6.8) | 7 (11.1) | 0.354 |
| **Norovirus GI** | 0 (0.0) | 1 (0.8) | >0.99 | 1 (1.1) | 0 (0.0) | >0.99 |
| **Norovirus GII** | 3 (11.5) | 9 (6.9) | 0.423 | 5 (5.7) | 7 (11.1) | 0.240 |
| **Enterovirus** | 1 (3.9) | 8 (6.2) | >0.99 | 4 (4.6) | 4 (6.4) | 0.720 |
| **CMV** | 0 (0.0) | 3 (2.3) | >0.99 | 2 (2.3) | 1 (1.6) | >0.99 |
| **Astrovirus** | 0 (0.0) | 3 (2.3) | >0.99 | 0 (0.0) | 2 (3.2) | 0.172 |
| **Rotavirus** | 0 (0.0) | 1 (0.8) | >0.99 | 0 (0.0) | 1 (1.6) | 0.417 |
| **Sapovirus** | 0 (0.0) | 0 (0.0) | - | 0 (0.0) | 0 (0.0) | - |
| **>1 virus detected** | 1 (3.9) | 5 (3.9) | >0.99 | 1 (1.1) | 5 (7.9) | 0.083 |
| **Bacteria** | 16 (61.5) | 62 (47.7) | 0.197 | 44 (50.0) | 32 (50.8) | 0.923 |
| **Shigella spp.** | 5 (19.2) | 24 (18.5) | >0.99 | 18 (20.5) | 9 (14.3) | 0.329 |
| **Salmonella spp.** | 2 (7.7) | 10 (7.7) | >0.99 | 6 (6.8) | 5 (7.9) | >0.99 |
| **C. difficile** | 2 (7.7) | 10 (7.7) | >0.99 | 6 (6.8) | 6 (9.5) | 0.544 |
| **Campylobacter** | 1 (3.9) | 10 (7.7) | 0.692 | 4 (4.6) | 7 (11.1) | 0.202 |
| **STEC** | 0 (0.0) | 0 (0.0) | - | 0 (0.0) | 0 (0.0) | - |
| **ETEC** | 1 (3.9) | 3 (2.3) | 0.522 | 3 (3.4) | 1 (1.6) | 0.641 |
| **EPEC** | 2 (7.7) | 8 (6.2) | 0.673 | 6 (6.8) | 5 (7.9) | >0.99 |
| **EAEC** | 2 (7.7) | 18 (13.9) | 0.531 | 10 (11.4) | 8 (12.7) | 0.803 |
| **O157** | 0 (0.0) | 3 (2.3) | >0.99 | 1 (1.1) | 2 (3.2) | 0.571 |
| **Plesiomonas** | 0 (0.0) | 1 (0.8) | >0.99 | 0 (0.0) | 1 (1.6) | 0.417 |
| **Helicobacter pylori** | 4 (15.4) | 8 (6.2) | 0.117 | 6 (6.8) | 6 (9.5) | 0.544 |
| **>1 bacteria detected** | 2 (7.7) | 24 (18.5) | 0.252 | 11 (12.5) | 13 (20.6) | 0.178 |
| **Parasite<sup>b</sup>** | 12 (46.2) | 59 (45.4) | >0.99 | 33 (37.5) | 37 (58.7) | **0.010** |
| **Cystoisospora** | 4 (15.4) | 24 (18.5) | >0.99 | 10 (11.4) | 17 (26.9) | **0.014** |
| **Cryptosporidium spp.** | 5 (19.2) | 21 (16.2) | 0.773 | 12 (13.6) | 14 (22.2) | 0.168 |
| **Blastocystis** | 4 (15.4) | 6 (4.6) | 0.063 | 8 (9.1) | 2 (3.2) | 0.195 |
| **Giardia spp.** | 1 (3.9) | 5 (3.9) | >0.99 | 4 (4.6) | 2 (3.2) | >0.99 |
| **Enterocytozoon spp.** | 0 (0.0) | 10 (7.7) | 0.215 | 4 (4.6) | 6 (9.5) | 0.321 |
| **Schistosoma** | 0 (0.0) | 4 (3.1) | >0.99 | 3 (3.4) | 1 (1.6) | 0.641 |
| **>1 parasite detected** | 2 (7.7) | 10 (7.7) | >0.99 | 7 (8.0) | 5 (7.9) | >0.99 |
| **Mixed infections** |  |  |  |  |  |  |
| **Virus-bacteria** | 3 (11.5) | 20 (15.4) | 0.768 | 11 (12.5) | 11 (17.5) | 0.394 |
| **Virus-parasite** | 2 (7.7) | 19 (14.6) | 0.531 | 8 (9.1) | 11 (17.5) | 0.126 |
| **Bacteria-**<br>**parasite** | 8 (30.8) | 34 (26.2) | 0.633 | 19 (21.6) | 22 (34.9) | 0.069 |

<sup>a</sup> ART status known for 156 and cotrimoxazole use known for 151 of 164 HIV-infected cases; <sup>b</sup> The following parasites were screened for but not detected: *E. histolytica*, *Strongyloides* spp., *Cyclospora*, *Hymenolepis*, *Ascaris*, *Taenia*, *Trichuris*, *Ancylostoma*, *Enterobius*, *Necator*, *Dientamoeba*.

